# A Model for the Testing and Tracing Needed to Suppress COVID-19

**DOI:** 10.1101/2020.06.02.20120568

**Authors:** Victor Wang

## Abstract

This paper presents the first analytical model for calculating how many tests and tracing needed to suppress COVID-19 transmission. The number of people needs to be tested daily is given by:

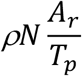

Where

*N* is the size of the population in consideration

*A*_*r*_ is the attack rate at any given time

*T*_*p*_ is the test-positive rate

ρ is the percentage of infectious people that have to be detected per day. To make the effective reproduction number *R*_*e*_ below 1, *ρ* must satisfy the following equation:

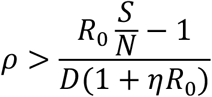

Where

*R*_*0*_ is the basic reproduction number,

S/N is the percentage of the susceptible population over the entire population,

*D* is the length of the infectious period, and

*η* is the percentage of close contacts that have to be traced.

This model provides insights and guidance to deploy the testing and tracing resources optimally. An Excel model is attached to facilitate easy calculation of the number of tests and tracing needed. This model is also applicable to any infectious disease that can be suppressed by testing and tracing.

## Introduction

As countries reopen their societies and economies from the COVID-19 lockdowns, the biggest concern is whether we will have new waves of infections. Before we have effective drugs and vaccines, the only measures we can rely on to suppress the transmission are social distancing such as wearing masks, testing to detect the infectious people to isolate them, and tracing their close contacts to quarantine them. This paper presents a simple mathematical model for the amount of testing and tracing needed to suppress the transmission, under the condition that certain social-distancing measures are in place, such as wearing masks in public.

The critical parameter of any infectious disease is its basic reproduction number *R*_*0*_, the average number of people can be infected by an infectious person during his entire infectious period, without any intervention or mitigation measures. With various intervention and mitigation measures in place, the reproduction number changes, the literature often denote *R*_*t*_ as the reproduction number at a given time *t*. The literature further defines *R*_*e*_ as the effective reproduction number taking into consideration of the effects of all the measures that can be taken, including social distancing, testing, and tracing with isolation. The goal of suppressing the transmission is to make *R*_*e*_ < 1.

One of the most widely used math models for infectious diseases is the SEIR model*(1,2)*. As depicted in Fig. 1, the total population in consideration is divided into four distinctive compartments, denoted as *S*-susceptible, *E*-exposed (those contracted the disease but not yet infectious), *I-*infectious, and *R*-removed.

**Fig. 1.**
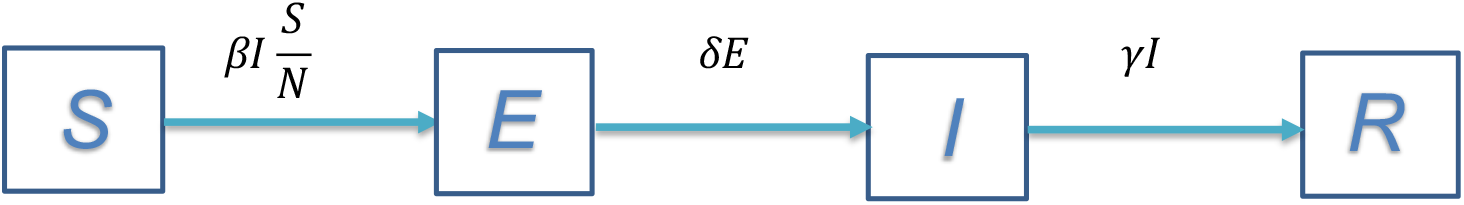
The SEIR compartment model for infectious diseases

In Fig. 1:

*S* – the size of the population of susceptible.

*E* – the size of the population exposed to the disease and infected but not infectious yet.

*I* – the size of the infectious population.

*R* – the size of the population removed from the infectious population *I*, including the recovered. All of the above four variables change over time as an infectious disease progresses.

*N* – the size of the entire population in consideration where *N = S + E + I + R*, which is a constant if without considering natural births and deaths, and the deaths due to the disease are negligible compared with *N*.

*β* -- the average daily number^1^ of people to be infected by an infectious person, when *S=N*.

*δ* -- the latent rate, 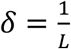, where *L* is the average length of time from being exposed to being infectious^2^.

*γ* -- the remove rate, 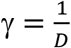, where *D* is the average length of the infectious period.

This model makes the following assumptions:

It ignores the effects of both natural births and deaths and the deaths due to the disease in consideration.

i. It defines “exposed” as those who already contracted the disease but not infectious yet, i.e., they are in the latent period.
ii. The “removed” are neither infectious nor subject to another infection, whether recovered clinically or not.

Since *β* is the daily number of people infected by an infectious person, the transfer rate from *S* to *E* is 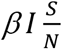. Note, even though each infectious person can infect *β* persons daily when *S=N*, as the population of *S* shrinks, fewer people can be infected, thus the daily number of new infected is scaled by the factor of 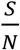. The transfer rate from *E* to *I* is reversely proportional to the length of the latent period *L*. Similarly, the transfer rate from *I* to *R* is reversely proportional to the length of the infectious period *D*.

With the above transfer rates between the compartments, we can have the following set of differential equations:

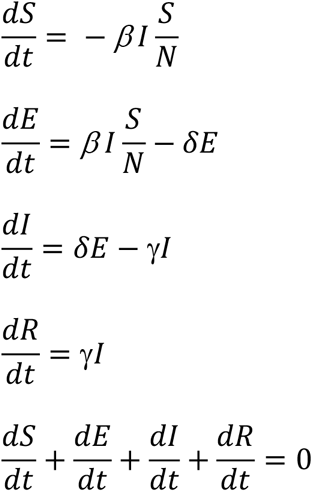

Given the above set of equations, with a set of initial conditions, we can have the populations’ size at any given time *t*. However, what we are most interested in here is whether the infected population will increase or decrease. The rate of change of the infected population is:

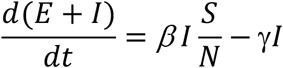

To suppress the transmission, we need the population of *E* + *I* to decrease over time, that is,

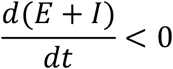

That is,

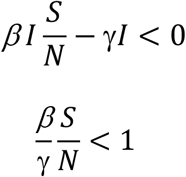

Since 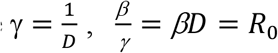, we have

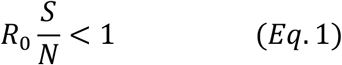

As an infectious disease progresses, *S* keeps decreasing, when 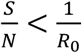, *Eq*. 1 is satisfied; thus, the disease will die out. The literature refers to such a scenario as the population reaches the “herd immunity”.

### The Modified SEIR Model Considering Testing and Tracing

Without intervention measures, the infected population can only decrease as people recover. The effects of testing and tracing are to artificially decrease the population of the infected by detecting and tracing then isolating them so that they can no longer infect others. The SEIR model depicted in Fig. 1 can be modified considering the effects of testing and tracing, as shown in Fig. 2

**Fig. 2.**
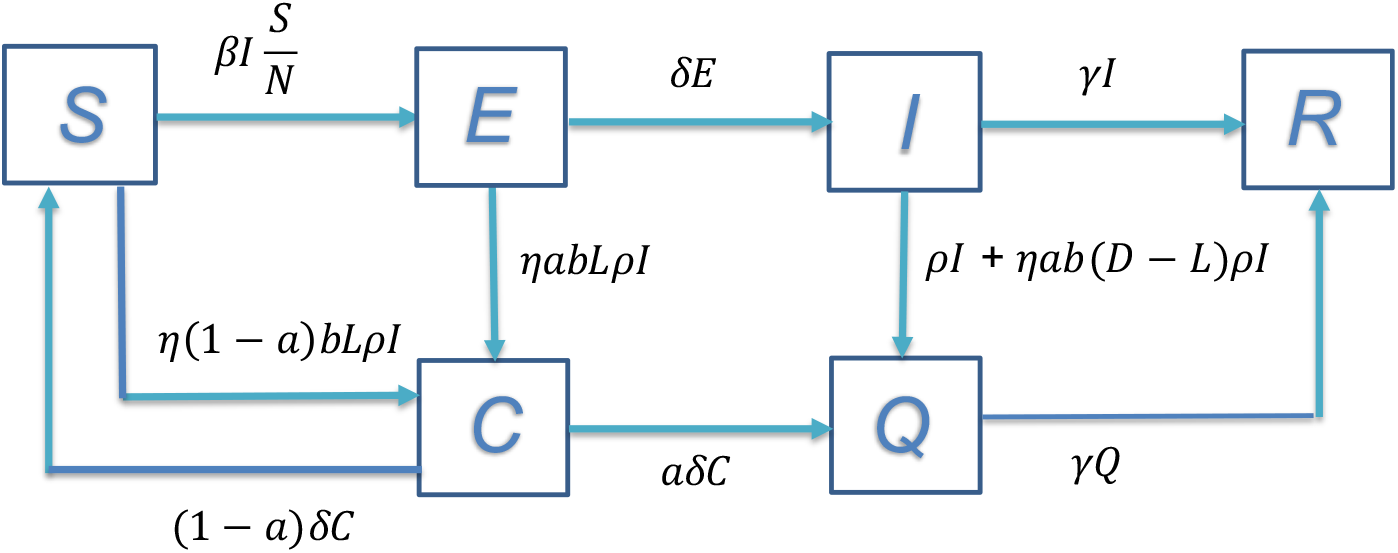
The Modified SEIR model with Testing and Tracing

Note here we add two more compartments:

*C* – the number of the close contacts traced and quarantined.

*Q* – the number of infectious people who are detected or traced and isolated.

We have:

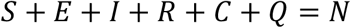

We further define *β* = *ab*, where:

*a* – the probability of a close contact to be infected by an infectious person.

*b* – the average number of close contacts an infectious person makes daily.

For measuring quantities of testing and tracing, we need to add two more variables as below:

*ρ* – the percentage of infectious people that are detected daily.

*η* – the percentage of close contacts that are traced daily.

By definition, the number of infectious people detected daily is *ρI*, the transfer rate from *I* to *Q* due to the detection is:

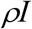

The number of close contacts made by each infectious person daily is *b*, then the number of close contacts made daily by the infectious people detected is *bρI*; thus the total number of close contacts made by these infectious people during their entire infectious period *D* days is *bDρI*. Therefore, the total number of close contacts that are traced daily is:

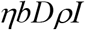

The close contacts can be in any of the three boxes *S, E*, and *I*, depending on when they had close contact with an infectious person. Those who had close contact with an infectious person within the *L* days are not yet infectious thus in either box *S* or *E*, while those who had close contact more than *L* days ago become infectious thus are in box *I*^*2*^. Now we have the following transfer rates due to the tracing:

From box *E* to *C*:

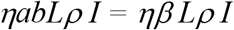

From box *S* to *C*:

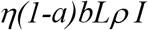

From box *I* to *Q* (in addition to the transfer rate due to the detection) is^4^:

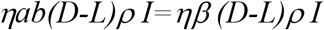

We assume the infectious people in box *Q* are removed at the same rate as those in box *I*, then the transfer rate from box *Q* to *R* is:

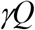

In box *C*, where there are both infected and non-infected. Those who are infected start to become infectious after their latent period, thus the transfer rate from box *C* to *Q* is:

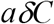

While the transfer rate from box *C* back to *S* is

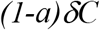

With the above transfer rates among the compartments established, we can have the following set of differential equations:

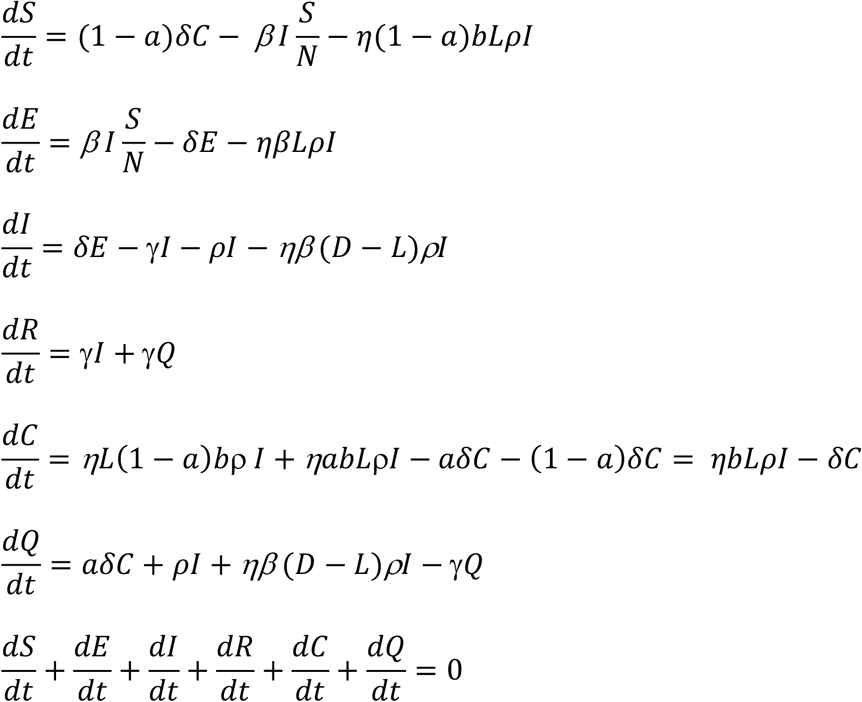

Given the above set of equations, with a set of initial conditions, we can have the populations’ sizes at any given time *t*, at least numerically. Again, here we focus on the changes in the infected population. Since *E+I* is the total population of infected, to suppress the transmission, we need *E* + *I* to decrease, that is,

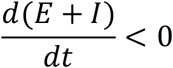

We have:

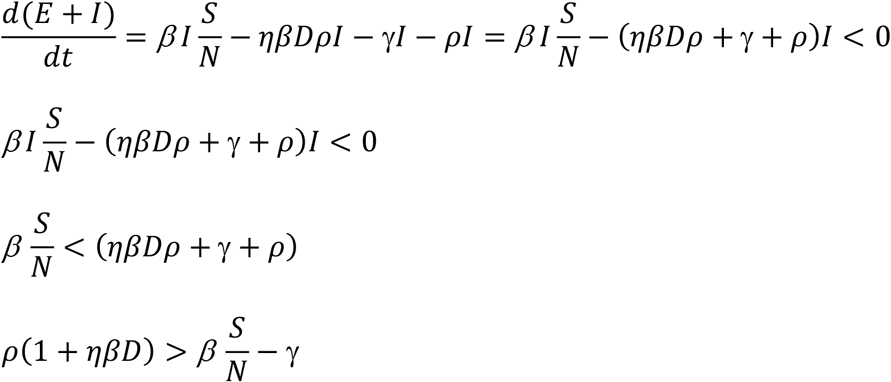

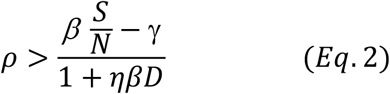

Since 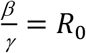, dividing both nominator and denominator of *Eq*. 2 with *γ*, we have:

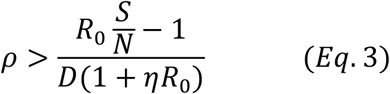

An equivalent form of Eq. 3 is:

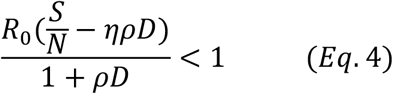

Note the left side of the *Eq*. 4 is nothing but the effective reproduction number *R*_*e*_, taking into the consideration of the effects of both testing and tracing. That is, if *Eq*. 3 holds, we have:

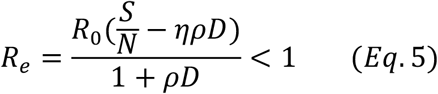

### How Much Testing and Tracing Needed in Practice ?

*Eq*. 4 defines the relationship between the daily testing ratio *ρ* and the daily tracing ratio *η* for various reproduction numbers *R*_0_. For COVID-19, authors in*(5)* quoted 12 studies of the basic reproduction number, mostly from China before any intervention measures were taken, and come up with the average *R*_*0*_ = 3.28 and median *R*_*0*_ = 2.79. Another study involving a large number of cases in Wuhan, China, shows that average *R*_*0*_ = 3.25 from January 1 to January 23, 2020, before Wuhan was locked down*(6)*.

When *R*_*0*_ is as high as 3.28, testing and tracing alone will not be able to suppress the transmission, we have to resort to additional measures such as social distancing. Among various social distancing measures, wearing masks in public has been proven effective. Research done at Hong Kong University shows that surgical masks can effectively reduce the transmission of various viruses*(7)*. The research provided data for the reduction of virus transmission in six different scenarios: transmitting droplets larger than 5 μm and aerosols smaller than 5 μm for coronavirus, influenza virus, and rhinovirus, respectively. Averaging over the six scenarios, wearing masks can reduce the transmission rate by 63%.

If we consider only wearing masks in social distancing and ignore the effects of other measures such as washing hands frequently and keeping physical distances, the reproduction number with social distancing, denoted as *R*_*s*_, can be 3.28 × 37% = 1.23. Note *R*_*s*_ is determined only by the effects of social-distance measures in place. In the following discussion, we assume the social-distance measures remain the same over time so that *R*_*s*_ does not change with time.

Note that *R*_*0*_ = 3.28 is among the highest values observed so far, mostly derived from the studies in large metropolitan areas like Wuhan in China, where population density is among the highest in the world. In the US, however, much lower *R*_*0*_ has been observed as indicated in the websites https://rt.live/ and https://covid19-projections.com/infections-tracker/. The websites quoted that the average *R*_*0*_ in the US is 2.22 before any intervention measure was taken. In all the states, only New York had *R*_*0*_ =3.66, while all other states had *R*_*0*_ lower than 2.5.

When we calculate the amount of testing and tracing needed, we need to consider the effects of social-distancing measures already in place. Thus, we need to use the reproduction number with social distancing *R*_*s*_ rather than the basic reproduction number *R*_*0*_. In the following discussion, Replacing *R*_*0*_ with *R*_*s*_ in *Eq*. 3, we have:

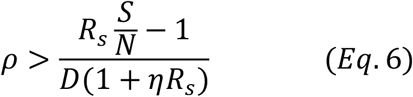

Since *S* changes over time, so too 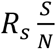 which is the reproduction number at time *t*, as defined in*(8)*:

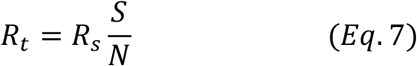

Similarly, *Eq*. 6 can be represented as:

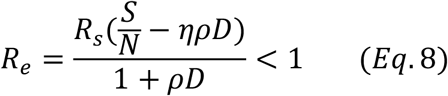

Now *R*_*e*_ is the effective reproduction number taking into consideration of the effects not only testing and tracing but also that of social distancing. It is clear here that *R*_*e*_ changes with time since *R*_*t*_ (or equivalently 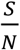) changes with time.

Our model depends on the value of *D* which is the average length of the infectious period. To suppress COVID-19 transmission, it is critical to understand when infected people start to shed virus, the intensity of the virus shedding over the infectious period, and how long an infected person can be infectious. Studies show that the median incubation period is 5.1 - 6.4 days*(9,10)*. A recent study by Singapore NCID shows that the infectious period of symptomatic individuals may begin around 2 days before the symptom onset and persists for about 7 - 10 days after the symptom onset*(11)*. Virus shedding was found highest in the first 2-4 days from symptom onset and remain high during the first week of symptoms*(12)*. In most cases, the virus could not be detected after 21 days from the symptom onset*(13)*. Another study shows that the viral load of severe cases is 60 times higher than the mild cases, 90% mild cases cleared virus after 10 days from symptom onset, but all severe cases not*(14)*. Active viral replication drops quickly after the first week, and a viable virus was not found after the second week of illness despite the persistence of PCR (Polymerase Chain Reaction) detection of RNA*(11,15)*.

Note that the shorter the *D* is for a given *R*_*s*_, the faster the disease spreads, thus harder to control. From the studies so far, the COVID-19 infectious period is found between 10-20 days, with 10-days being the most severe condition, hence the most conservative assumption to use in modeling. In the following two plots, we use *D*=10 days, and 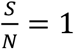 as a conservative bound.

Fig. 3 is the plot of daily detection ratio vs. daily tracing ratio, for *R*_*s*_ = 1.1, *R*_*s*_ = 1.2, and *R*_*s*_ =1.3, respectively. For *R*_*s*_ = 1.1, the maximum amount of detection we need to suppress the transmission is to catch 1% of the infectious population per day, assuming we do not do any tracing. If we can trace 100% of close contacts, we only need to detect 0.5% of the infectious population daily. For the scenario of *R*_*s*_ = 1.2, if we can trace only 50% of the close contacts, we would need to detect 1.3% of the infectious population daily.

**Fig. 3.**
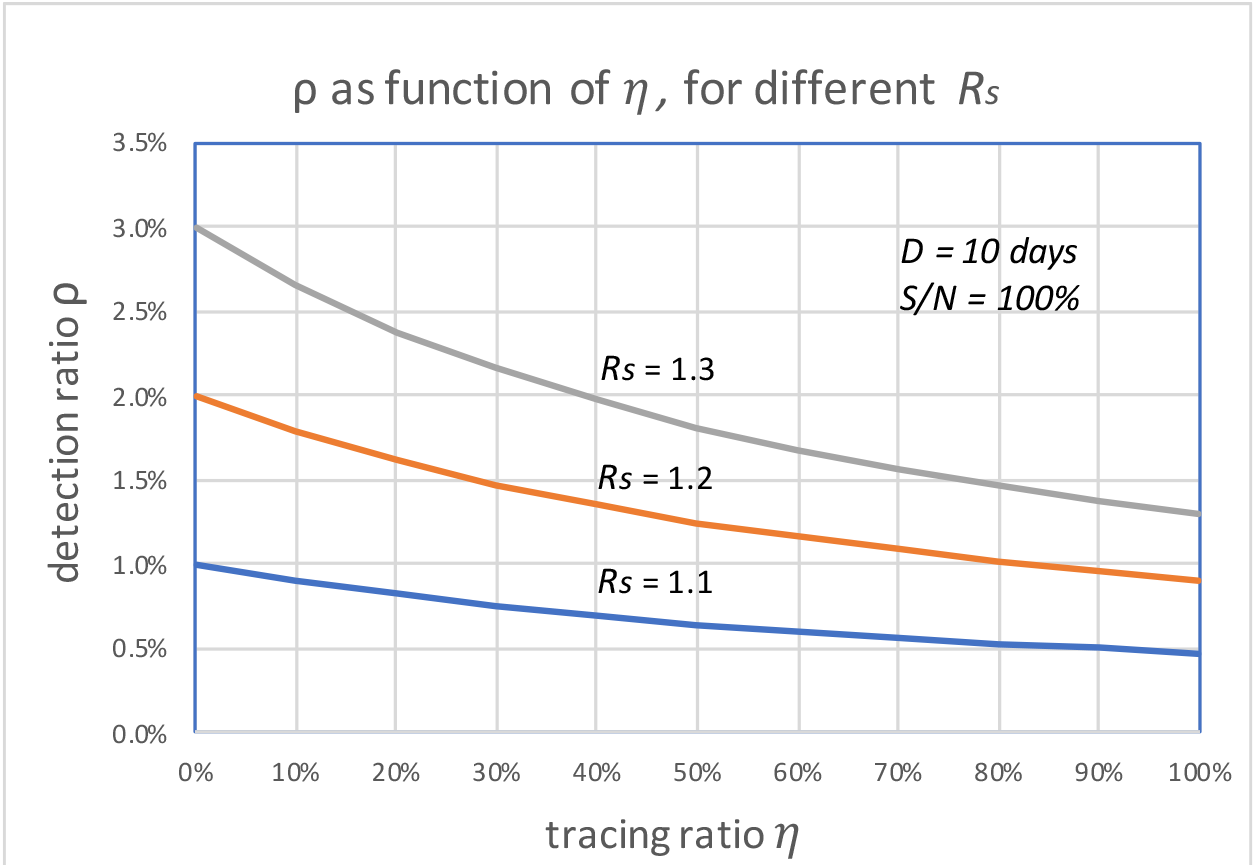
The daily detection ratio vs. daily tracing ratio, for different R_s_

Fig. 4 depicts the detection ratio as function of *R*_*s*_ for three different tracing ratios *η* = 25%, *η* = 50%, and *η* = 75%, respectively. We notice that as *R*_*s*_ increases, the quantity of detection required increases quickly. When *R*_*s*_ = 2.0, if we can trace 50% of close contacts, we would need to detect 5% of infectious people per day. It is also worth noting that when *R*_*s*_ is small, e.g., close to 1.0, tracing does not make as much a difference as when *R*_*s*_ is larger. For the given costs of the detection and the tracing, we can find the optimal quantities of testing and tracing to minimize the total cost. For example, as the testing becomes more available and costs lower, if tracing is too hard to do, we may choose to do more testing to achieve the same goal.

**Fig. 4.**
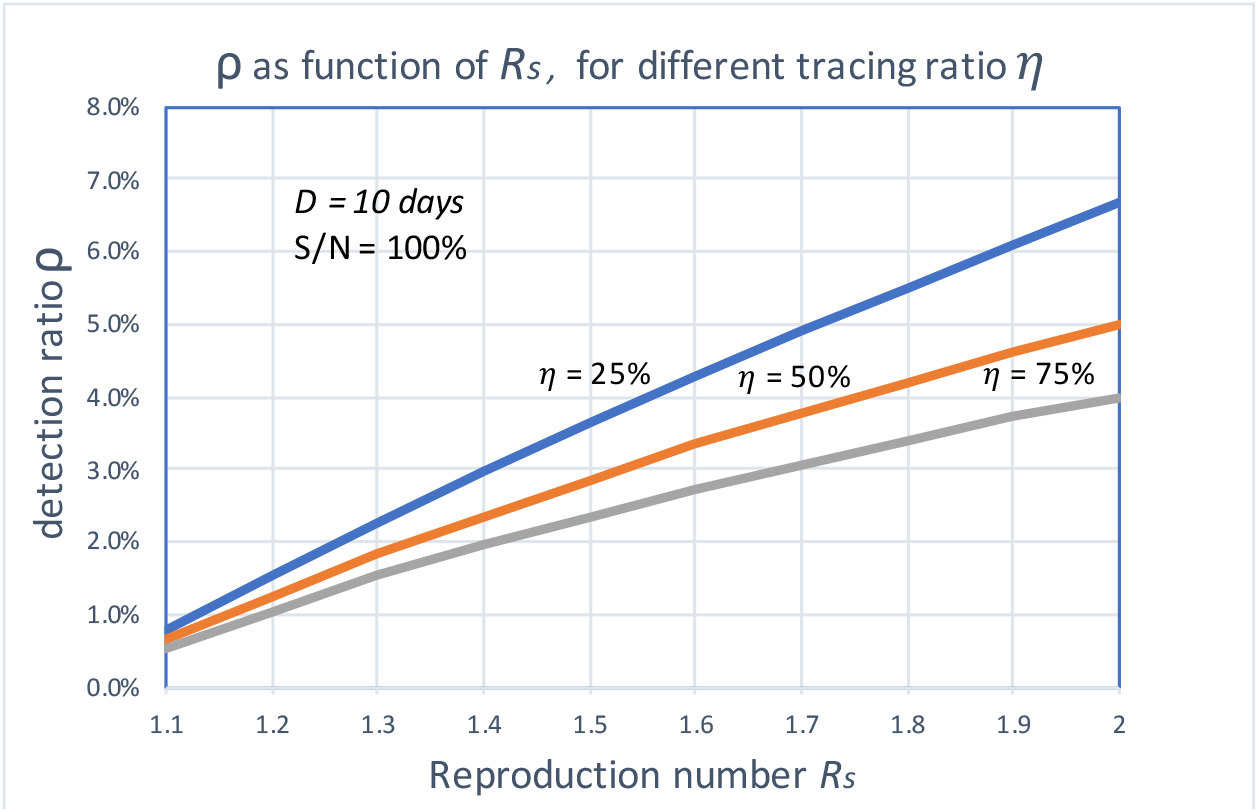
The daily detection ratio as a function of R_s_ for different tracing ratios

In both Fig. 3 and Fig. 4, we assume 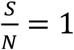 that is a conservative bound, since as the disease progresses, *S* keeps decreasing. As of May 28, 2020, both infection and seroconversion^5^ Surveys have been conducted in many places in the world. The UK government published data that as of May 24, 2020, 6.78% (95% confidence interval: 5.21% to 8.64%) of individuals from whom blood samples were taken tested positive for antibodies to the coronavirus*(16)*. The survey results from studies in Spain and France indicate that 5.0% and 4.4% of their populations have ever contracted coronavirus*(17)*. In many metropolitan areas of the world, the seroconversion rate is much higher, for example, that of Boston is 10%*(18)*, Madrid 11%*(19)*, Moscow 14%*(20)*, London 17%*(19)*, and New York City 19.9% while New York State 12.3% *(21)*. With the above data, as of May 28, 2020, it seemed that for many European countries, the nationwide seroconversion rate had reached about 5%, while in many metropolitan areas, it has reached about 15%. That implies the 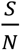 has been at least reduced to 95% and 85%^6^, respectively. In the following two Fig.s, we plot the testing and tracing needed for these three values of 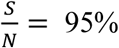 90%, and 85%, respectively.

In Fig. 5, we can see that the value of 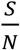 can significantly affect the outcome. For example, for *R*_*s*_ =1.2 and when 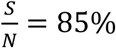, if we can trace 50% of the close contacts, we need to detect only 0.13% of infectious people per day. Compared with when 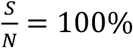 (as shown in Fig. 3), where we need to detect 1.3% of the infectious population per day, 0.13% is only 1/10 of the testing required.

**Fig. 5.**
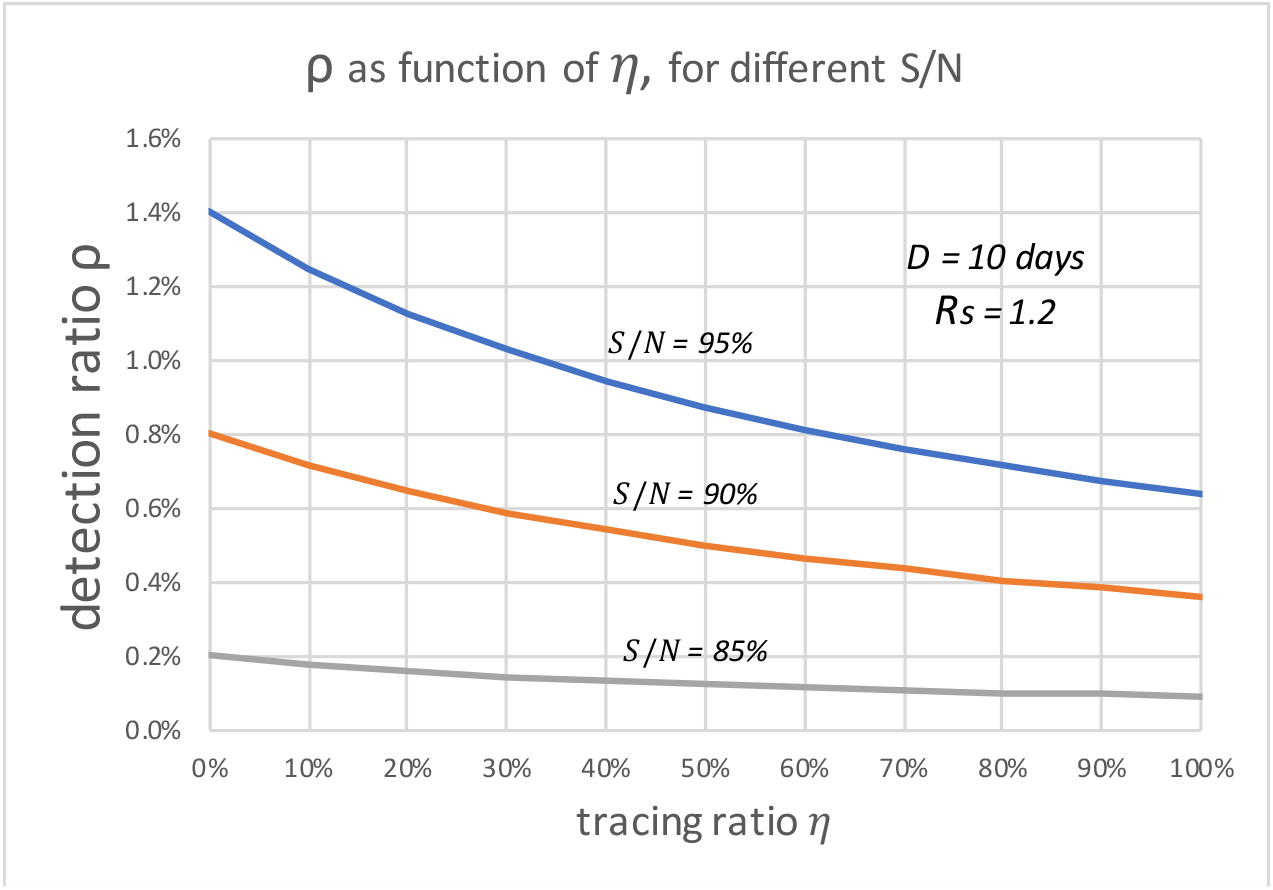
The daily detection ratio vs. daily tracing ratio, when Rs=1.2, for different S/N

In Fig. 6, we can see that for *R*_*s*_ < 1.1 and 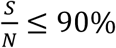

**Fig. 6.**
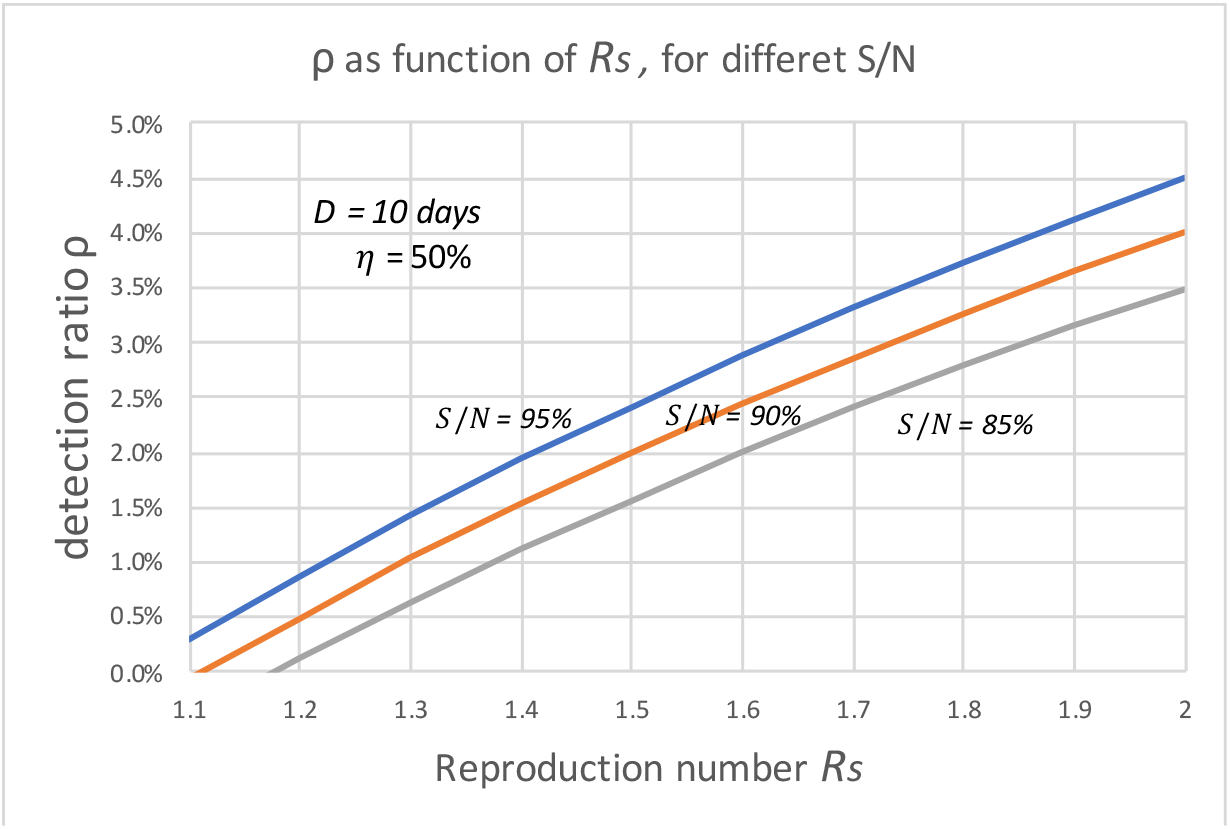
The daily detection ratio vs. Rs, when η = 50%, for different S/N

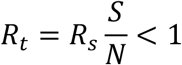

That means the disease will die out without needing to do any testing and tracing.

## Discussion on Implementations

The first question we have in implementing the testing and tracing is: how do we know the detection and tracing ratios achieved?

Let us first discuss the detection ratio. One way to estimate the detection ratio is to do a large-scale random infection survey over the population in consideration (except those recovered and those tested positive with antibodies) to come up with the attack rate (the percentage of infectious at any given time). With the attack rate established, we can estimate the total number of infectious people in the population from which we can calculate the detection ratio.

To estimate the tracing ratio, we can count those close contracts traced who eventually become infectious for per infector, denoted as *I*_*ct*_. Then the tracing ratio is

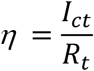

While we can know the detection ratio right after the daily detection is done, we may have to wait for a few days (the average length of the incubation period of COVID-19 is 5.1 days) before we can count in all the close contacts who become infectious (assume we only test them when they have symptoms unless we test them every day).

The second question is how to calculate the number of people to be tested per day. For example, if our goal is to detect 1% of the infectious people per day. We can use a brute-force approach to accomplish such a goal: to test 1% of the entire population (except for those who tested positive or with antibodies). This approach assumes the infectious people are randomly distributed in the entire population, which is certainly not true; thus, we can do a better job than the brute-force approach. With a given testing capacity, we can test the target groups in the population with the highest risk of contracting the virus first.

In general, we need to know both the attack rate and the test-positive rate. Let us define as:

*A*_*r*_ – Attack rate, the percentage of the infectious people in the entire population at a given time.

*T*_*p*_ – Test-positive rate, the percentage of positive results in the target groups to be tested

Let us further define:

*N*_*t*_ – Number of people need to be tested per day to satisfy required *ρ* for a given *η*.

We know:

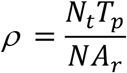

Where *N* is the size of the total population in consideration, thus we have:

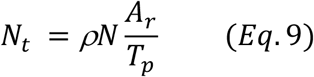

From *Eq*. 9, we immediately have the percentage of the population needs to be tested per day:

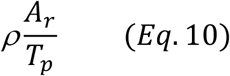

Note here that the brute-force approach assumes that *T*_*p*_= *A*_*r*._ It is clear here that the ratio of 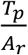 represents the concentration of the infectious people in the target groups tested.

For example, we should test anyone who has suspected symptoms and encourage everyone who suspects themselves having symptoms to come to a test. The survey*(16)* in the UK showed that the test-positive rate of those with specific symptoms (reporting having cough, fever, and loss of taste or smell) is 6.7%, that of people without symptoms is only 0.4%, while that of those with any one of the symptoms is 2.6%. In the same survey, the attack rate was given as 0.43%; if we apply all the testing capacity to those with specific symptoms, we can detect 1% of infectious people with testing only (0.43/6.7)x1%= 0.06% of the population. If we apply the testing capacity to those with any one of the symptoms, we need only to test (0.43/2.6)x1% = 0.17% of the population to detect 1% of infectious people. These represent much improvement over the brute-force approach.

In practice, we need to categorize the entire population into groups according to their risk levels. For example, in the UK, those who reported working in patient-facing healthcare workers have 1.73% tested positive. In comparison, the percentage of people reporting not working in these types of roles test-positive rate was much lower at 0.38%*(16)*. We need to do more of such surveys for different groups of the population. We can detect more infectious people with the minimum testing capacity if we apply limited testing capacity to the highest-risk groups first.

The third issue in implementation is how to estimate *R*_*s*_ of the region of the consideration, or equivalently how to estimate *R*_*t*_ (since we can estimate 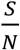 by doing a seroconversion survey, we can have *R*_*s*_ when we know *R*_*t*_ from *Eq*. 7). The estimation of *R*_*t*_ has been discussed extensively in the literature, and tools are also provided(*8)*(*22-25*). Websites such as https://rt.live/ also provide the real-time estimation of the *R*_*t*_ for all 50 states of the US.

## Applications of the Model

The model developed in this paper is the most appropriate for situations where there are already a large number of infected people spread out in the community, like in the US and many other European countries. For countries like China where after a period of strict lockdown, the new cases come to almost zero, or Taiwan, where they did excellent job suppressed the infections from very beginning to a minimal number*(26)*, they do not need to do a large amount of testing to detect the infectious people from the population. Given there are so few new cases in these places, all they need to do is pay close attention to any new cases, including screening out all imported cases and tracing all close contacts of these new cases.

This model should apply to any relatively isolated communities, such as a country, a state, a metropolitan area, or even a remote town. As many private employers are taking the testing into their own hands, this model may or may not apply to a company where their employees have frequent interactions with members outside the company.

An excel model is provided as an appendix of this paper to facilitate easy calculation. The excel model used the data from California on July 20^th^, 2020. Users can enter their own data in the highlighted cells to calculate the number of people that need to be tested per day to suppress COVID-19 transmission. For questions using the excel model, please correspond to the author at.

## Conclusion

With social-distancing measures in place, the reproduction number of COVID-19 has been significantly reduced in all places. It might have been reduced to below 1 in places where more disciplined social-distancing is practiced, such as in Japan. In such places, extensive testing and tracing may not be needed. However, in other places of the world, depending on the scale and degree of the reopening, the reproduction number may come back to be higher than 1. In these places, we can rely on conducting a reasonable amount of testing and tracing to suppress the new waves of infections, before any effective drugs and vaccines become available. The model developed here can provide quantitative guidance on how to deploy the testing and tracing resources optimally. Last but not least, this model applies to any infectious disease that can be suppressed by testing and tracing.

## Data Availability

N/A

## Appendix

**An Excel Model for Calculating the Number of People to be Tested Daily**

## Footnotes

1 Note here we use “daily number” to make it easy to understand. In rigorous math terms, the “daily number” should be the number occurring in unit time. We use “daily number” or “number per day” throughout the text without further noting.

2 Here we assume the exposure time is the same as the infection time although in general, that may not always be true. Note also that the latent period is not the same as the incubation period unless the patients become infectious exactly when they start to have symptoms*(3,4)* In any case, these differences do not affect our final result since it does not depend on this parameter *δ*.

3 Here we assume the length of the infectious period *D* is equal or longer than the length of the latent period *L*, which is the case for COVID-19.

4 We assume testing every close contact as soon as they are traced so we know who is infectious to be isolated.

5 Measuring the percentage of the population having developed antibodies from the past infections of SARS-CoV-2 virus.

6 We assume these seroconversion test results are accurate and ignore the size of the population who are infected but have yet developed antibodies.

## References

1. Fred Brauer, Carlos Castillo-Chavez, and Zhilan Feng. Mathematical Models in Epidemiology (Texts in Applied Mathematics Book 69). Springer 2019

2. Compartmental models in epidemiology. Wikipedia. https://en.wikipedia.org/wiki/Compartmental_models_in_epidemiology

3. Latent period (epidemiology). Wikipedia. https://en.wikipedia.org/wiki/Latent_period_(epidemiology)

4. Arnold Bosman. Incubation period, Latent period and Generation time. Field of Epidemiology Manual Wiki. https://wiki.ecdc.europa.eu/fem/Pages/Incubationperiod,LatentperiodandGenerationtime..aspx

5. Ying Liu, Albert A. Gayle, Annelies Wilder-Smith, and Joacim Rocklöv. The reproductive number of COVID-19 is higher compared to SARS coronavirus. Journal of Travel Medicine, Volume 27, Issue 2, March 2020. https://academic.oup.com/jtm/article/27/2/taaa021/5735319

6. Pan A, Liu L, Wang C, et al. Association of public health interventions with the epidemiology of the COVID-19 outbreak in Wuhan, China. JAMA. Published online April 10, 2020. doi:10.1001/jama.2020.6130

7. Leung N, Chu D, Shiu E, Chan KH, McDevitt J, Hau B et al. Respiratory virus shedding in exhaled breath and efficacy of face masks. Nature Medicine, April 3 2020. https://www.nature.com/articles/s41591-020-0843-2

8. Luís M. A. Bettencourt, Ruy M. Ribeiro. Real Time Bayesian Estimation of the Epidemic Potential of Emerging Infectious Diseases. PLOS ONE. May 14, 2008 https://doi.org/10.1371/journal.pone.0002185

9. Lauer S, Grantz K, Bi Q, Jones F. The Incubation Period of Coronavirus Disease 2019 (COVID-19) From Publicly Reported Confirmed Cases: Estimation and Application. March 10, 2020. Annals of Internal Medicine https://www.acpjournals.org/doi/10.7326/M20-0504

10. Backer JA, Klingkenberg D, Wallinga J. Incubation period of 2019 novel coronavirus (2019-nCoV) infections among travelers from Wuhan, China, 20-28 January 2020. Euro Surveill. 2020 Feb;25(5). https://www.ncbi.nlm.nih.gov/pubmed/32046819?dopt=Abstract

11. Position Statement from the National Centre for Infectious Diseases and the Chapter of Infectious Disease Physicians. Academy of Medicine, Singapore. May 23, 2020. Singapore study on infectious period

12. Wölfel R, Corman V, Guggemos W, Seilmaler M, Zange S, Müller M, et al. Virological assessment of hospitalized cases of coronavirus disease 2019. Nature 581, 465–469 (2020), April 1, 2020. https://www.nature.com/articles/s41586-020-2196-x

13. Zou L, Ruan F, Huang M, Liang L, Huang H, Hong Z. et al.. SARS-CoV-2 Viral Load in Upper Respiratory Specimens of Infected Patients. N. Engl. J. Med. 382, 1177–1179 (2020). https://www.nejm.org/doi/10.1056/NEJMc2001737

14. Liu Y, Yan L, Wan L, Xiang T, Le A, Liu J et al. Viral dynamics in mild and severe cases of COVID-19. Lancet, March 19, 2020. https://www.thelancet.com/journals/laninf/article/PIIS1473-3099(20)30232-2/fulltext

15. He X, Lau E, Wu P, Deng X, Wang J, Hao X et al. Temporary Dynamics in Viral Shedding and Transmissibility of COVID-19. Nat. Med. April 15 2020. 2020 May; 26(5):672:675 https://pubmed.ncbi.nlm.nih.gov/32296168/

16. UK. Coronavirus (COVID-19) infection survey Pilot: May 28. Office for National Statistics. The link to the survey

17. Jacqui Wise. Covid-19: Surveys indicate low infection level in community. BMJ 2020;369:m1992 doi: 10.1136/bmj.m1992 (Published May 18 2020)

18. https://www.boston.com/news/coronavirus/2020/05/15/boston-coronavirus-antibody-testing-results

19. The Conversation. Herd immunity in Europe – are we close? https://theconversation.com/herd-immunity-in-europe-are-we-close-139253

20. 14% of Healthy Russians Have Coronavirus Antibodies, Private Lab Says https://www.themoscowtimes.com/2020/05/22/14-of-healthy-russians-have-coronavirus-antibodies-private-lab-says-a70353

21. https://www.governor.ny.gov/news/amid-ongoing-covid-19-pandemic-governor-cuomo-announces-results-completed-antibody-testing

22. A. Cori, N.M. Ferguson, C. Fraser, and S. Cauchemez. A New Framework and Software to Estimate Time-Varying Reproduction Numbers During Epidemics [link] 2013. American Journal of Epidemiology, Vol 178(9), pp. 1505–1512. DOI: 10.1093/aje/kwt133

23. Burr T1, Chowell G. “The reproduction number R(t) in structured and nonstructured populations.” Math Biosci Eng. 2009 Apr;6(2):239–59, https://www.ncbi.nlm.nih.gov/pubmed/19364151

24. Thompson R, Stockwin J, Gaalen R, Polonsky J, Kamva Z, Demarsh P, et. al., “Improved inference of time-varying reproduction numbers during infectious disease outbreaks” Epidemics, Volume 29, December 2019, https://www.sciencedirect.com/science/article/pii/S1755436519300350

25. Zhao S, Cao P, Gao D, Zhuang Z, Cai Y, Ran J, et al., “Serial interval in determining the estimation of reproduction number of the novel coronavirus disease (COVID-19) during the early outbreak”, Journal of Travel Medicine. taaa033, https://doi.org/10.1093/jtm/taaa033

26. Wang J, Ng C, Brook R. Response to COVID-19 in Taiwan Big Data Analytics, New Technology, and Proactive Testing. Journal of American Medical Association, March 3, 2020. https://jamanetwork.com/journals/jama/fullarticle/2762689

